# Development and validation of a 5-year risk model using mammogram risk scores generated from screening digital breast tomosynthesis

**DOI:** 10.1101/2024.09.17.24313569

**Authors:** Shu Jiang, Debbie L. Bennett, Graham A. Colditz

**Affiliations:** Division of Public Health Sciences, Department of Surgery (S.J., G.A.C.), Department of Radiology (D.L.B.), Washington University School of Medicine in St. Louis, St. Louis, MO

## Abstract

Screening digital breast tomosynthesis (DBT) aims to identify breast cancer early when treatment is most effective leading to reduced mortality. In addition to early detection, the information contained within DBT images may also inform subsequent risk stratification and guide risk-reducing management. We obtained a 5-year area under the curve (AUC) = 0.78 (95% confidence interval (CI) = 0.75 – 0.80) in the internal validation. The model validated in external data (n=6,553 women; AUC = 0.77 (95% CI, 0.74 – 0.80). There was no change in the AUC when age and BI-RADS density are added to the synthetic DBT image. The model significantly outperforms the Tyrer-Cuzick model (p<0.01). Our model extends risk prediction applications to synthetic DBT, provides 5-year risk estimates, and is readily calibrated to national risk strata for clinical translation and application in the setting of US risk management guidelines. The model could be implemented within any digital mammography program.

**One Sentence Summary:** We develop and externally validate a 5-year risk prediction model for breast cancer using synthetic digital breast tomosynthesis and demonstrate clinical utility by calibrating to the national risk strata.

## Introduction

To date, applications of radiomic data has largely focused on improved diagnosis at the time of screening. The widespread use of digital mammography supported this focus for both full field digital mammograms (FFDM) and digital breast tomosynthesis (DBT). Concurrent with diagnosis, breast cancer risk prediction has an increasing role in routine screening. Prediction models have moved from using demographic variables that approximate changes in the breast tissue to using digitized film mammograms, and now to digital images^1^ to predict long-term risk. Digital breast tomosynthesis (DBT) has been evaluated and internally validated to predict up to 2-year risk.^2^ Given that current guidelines in the US refer to 5-year risk as a guide for risk management, including chemoprevention, other risk reduction strategies,^3,4^ and additional screening,^5^ it is imperative that long-term risk be evaluated. Furthermore, these long-term prediction models must be evaluated in a diverse population to ensure generalizability.^6^

Digital breast tomosynthesis (DBT) was approved by FDA for all women in 2011.^7^ It improves the cancer detection rate on screening ^8^ and has demonstrated usefulness in screening and diagnostic settings.^9^ The uptake of tomosynthesis for breast screening has varied over time by insurance status and other population characteristics.^7,10^ It was approved by Medicare from 2015 but through 2017 women from underrepresented race/ethnic groups, lower education, income, and rural residences had been slower to access this technology.^10^ This is consistent with other advanced imaging technologies including MRI.^11^

Given the differential uptake over time and yet now widespread use, it is imperative that long-term risk of breast cancer also be evaluated. With sufficient time from implementation to identify cases of breast cancer in a broadly screened population, a prediction tool that works across populations is urgently needed for clinical translation. We draw on an American College of Radiology (ACR) accredited and designated comprehensive breast imaging center providing routine breast screening in a diverse population (27% Non-Hispanic Black women)^12^ to develop and apply a risk prediction model from the DBT screening images. For the first time, we illustrate the model performance in an external validation cohort with 46% of Non-Hispanic Black women.^13^ We use a 5-year risk horizon to assess prediction performance following DBT screening mammography and demonstrate clinical utility by calibrating to the national risk strata to better manage risk.

## Methods

### Analytic data set

The Joanne Knight Breast Health Cohort at Washington University (WashU cohort) is used as training data in this study with up to 10 years of mammograms.^14^ This cohort of women undergoing routine mammography screening in St. Louis includes over 10,000 women with 27% non-Hispanic Black women. ^14^ Incident breast cancers (invasive and in situ) are identified through record linkage to pathology and tumor registries. Eligibility included consent for follow-up and attending a routine screening visit. We excluded women whose entry examination resulted in diagnosis of breast cancer and those with a diagnosis within first 6 month of their mammogram. We began follow-up from the first DBT through December 2020. We identified 105 breast cancer cases excluding those with diagnosis in the first 6 month of their eligibility DBT among 5,066 women free from breast cancer at their first screening DBT. The flow of participants through the study is set out in supplementary Figure 1. All mammograms were uniformly processed on Hologic machines. On entry to the cohort, women self-reported breast cancer risk factors using established and validated measures.^15^ From these data we estimated the Tyrer-Cuzick model risk of breast cancer.^16,17^

### External validation cohort

The external validation data set is drawn from the Emory Breast Imaging Dataset (EMBED) with up to 8 years of follow-up.^13^ This cohort represents a 20% random sample with de-identified mammograms of diverse women undergoing screening or diagnostic mammograms at 4 hospitals (2 community, one large inner-city and one private academic hospital) from January 2013 through December 2020. Similar to the WashU cohort, we include women from their first DBT and excluded women diagnosed with breast cancer within the first 6 month since entry to cohort, and those who had a history of breast cancer prior to first DBT. We identified 101 pathology-confirmed breast cancer (including in situ cases) beyond 6 months of entry to cohort and 6,453 women (46.1% Non-Hispanic Black) who remained free from breast cancer during follow-up. Data included age, race, and time from initial digital screening mammogram to breast cancer diagnosis. DBT were performed all on Hologic machines.

### Statistical Analysis

The risk model using synthetic DBTs was developed in the WashU cohort of over 10,000 women. Our model takes the synthetic DBT mediolateral oblique (MLO) and craniocaudal (CC) images with the option to input clinical risk factors. The outputs of the model include a mammogram risk score (MRS), probability of 5-year breast cancer onset, and relative risk for each woman that can be used for risk calibration (see Figure 1).

Performance of the following models are examined: 1) age and BI-RADS density; 2) synthetic DBT mammogram only; 3) synthetic DBT plus age and BI-RADS density.

We performed both an internal validation and an external validation in assessing the prediction performance. The internal validation uses a 5-fold cross-validation and involves randomly partitioning women in the WashU cohort into 5 subsamples. The external Emory data are used solely for validation. The 95% confidence intervals were estimated using 5000 bootstraps.

The performance of the risk prediction model was assessed in terms of discrimination of risk stratification. We used the area under the ROC curve (AUC) in assessing the discrimination performance.^18^ We report the 5-year AUC from the first screening synthetic DBT. We generated a distribution plot to visualize the mammogram risk score (MRS) separation between women who subsequently develop breast cancer and women who remain cancer-free in the external validation cohort. The US SEER calibrated risk stratification by absolute age using MRS is also reported in the external validation. Additionally, we show calibration via predicted vs. observed 5-year risk in the external validation. Because prediction performance has varied in population groups, we stratified sub-analyses by race (Non-Hispanic Black vs. Non-Hispanic White), and density (dense vs. non-dense).

This prospective cohort study creation and follow-up was supported by WashU and the Breast Cancer Research Foundation. Ethical approval was obtained from the Institutional Review Board. Informed written consent was obtained for study participation. The Emory cohort de-identified data were shared following Institutional Review Board approval.^13^

## Data availability

Development data mammogram images and covariates at WashU - available with data use agreement

External validation data from Emory publicly available

https://github.com/Emory-HITI/EMBED_Open_Data

## Results

### Cohort characteristics

We draw on a cohort of 5,066 women consented for linkage to medical records, including pathology reports to study risk of breast cancer in a diverse population (WashU cohort). We developed the model in this diverse population that includes uninsured women and those covered through the Breast and Cervical Screening Program as well as other forms of insurance. The external validation (Emory cohort) is in a similar diverse population of 6,553 women.

Breast cancer risk factors were assessed at entry to cohort in this prospective study (Table 1). Mean age at entry was 54.6 for women who remained cancer free. 26.5 % of the population self-identified as Non-Hispanic Black women. Cases had denser BI-RADS (C and D) and had higher prevalence of family history of breast cancer (25.5% compared with women who remained cancer free (17.1%)). Comparable BI-RADS distribution and ethnic diversity in the Emory external validation cohort are reported in Table 1. The mean age at first DBT was 55.8 for women who remained cancer free and 46.1% are NHB. Cases of breast cancer were diagnosed consistently across all 5 years of follow-up.

### 5-year risk prediction performance

All results here exclude women who have developed breast cancer within the first 6 month of their screening mammogram. We first assessed a model using age and BI-RADS density on entry. We observed a 5-year AUC of 0.55 (95% CI, 0.53, 0.57) in both internal and external validation. We then assessed model with the synthetic DBT mammogram only. We observed a 5-year AUC of 0.78 (95% CI, 0.75 – 0.80) in the internal validation and 0.77 (95% CI, 0.74 – 0.80) when applied to the external validation data. There was no change in the AUC when age and BI-RADS density are added to the DBT image (Table 2).

While many demographic risk factor models are available, we chose to use the Tyrer-Cuzick (TC) model in our development cohort as a representative demographic model. Comparison with the TC model was done in the WashU cohort only due to the lack of risk factors in Emory public access data. We obtained a 5-year AUC of 0.56 (95% CI, 0.54, 0.58) using TC alone, and AUC of 0.78 (95% CI, 0.75, 0.80) when we added TC with the synthetic DBT (Supplementary Table 2). The model using the synthetic DBT statistically significantly outperforms the TC model (p<0.01).

### Synthetic DBT mammogram risk score and risk calibration

The distribution plot of synthetic DBT mammogram risk scores (MRS) in the external validation cohort shows good separation between women who developed breast cancer and those who remained cancer-free, consistent with the observed AUC (Figure 2A). The estimated odds ratio (OR) for 1 standard deviation increase in MRS was 4.2 (95% CI, 4.0, 4.4).

### Calibration to the population risk

The MRS is also used to calibrate to the US SEER 5-year expected incidence of breast cancer in the external validation cohort (Figure 2B). Using the SEER 5-year cut points, the high-risk category (4% or higher 5-year risk) includes 49% of breast cancer cases and 15% of the cancer-free women (Supplementary Table 3). The very-low-risk category (lower than 1% 5-year risk) includes 10% of the breast cancer cases diagnosed in the next in 5 years and 51% of the women who remained breast cancer-free (supplementary table 3). A risk ratio of 16.9 was observed when comparing the high-risk group (5-year risk > 4%) to the very-low-risk group (5-year risk < 0.3%) (Figure 2B).

### Calibration of observed and predicted risk

We present predicted vs. observed 5-year risk by decile with their 95% confidence interval in Supplementary Figure 2. It shows good calibration across all levels of risk. We did not observe any over or under estimation of risk across all deciles of risk.

### Sensitivity Analyses

We next assessed model performance predicting risk for varying time intervals after the current clinic visit. As seen in table 1, the diagnosis of breast cancer cases is well distributed across time from first DBT in both data sets. In the external validation data, performance was steady from AUC of 0.82 for 2-4-year risk and 0.80 for 5-year risk (Table 3).

Stratified sub-analyses of performance in the external validation data by race/ethnicity, BI-RADS breast density, are presented in Supplementary Table 1. Because breasts with high mammographic density hinder detection of tumors, we stratified women to dense (BI-RADS C/D) and non-dense (BI-RADS A/B). Among women with dense breasts at the first DBT, we obtained a 5-year risk AUC of 0.77 (95% CI, 0.73 - 0.82) in the external validation data. An AUC of 0.78 (95% CI, 0.73 - 0.83) was obtained for the non-dense subgroup. Additionally, we obtained a 5-year risk AUC of 0.78 (95% CI, 0.74 - 0.82) for Non-Hispanic White women and 0.75 (95% CI, 0.70 - 0.80) for Non-Hispanic Black women.

## Discussion

For the first time, we developed and externally validated the performance of a 5-year breast cancer risk prediction model using synthetic DBT images in a diverse external validation population that is 46% non-Hispanic Black women. The synthetic DBT image-based prediction model performed equally well in Non-Hispanic White and Non-Hispanic Black women and across women with dense and non-dense breasts. We extend prediction from synthetic DBT images to 5-year risk and reached a 5-year AUC of 0.77 in the diverse external validation cohort. This study shows feasibility for risk estimation from synthetic DBT images in routine breast screening services for long-term risk prediction consistent with current US risk management guidelines.

Risk stratification is an essential first step to either implementing risk reduction strategies among high-risk women,^4,19^ or moving to precision prevention where screening modality and frequency might vary as well as the use of lifestyle and chemoprevention approaches to reduce risk and improve risk benefit outcomes for the screened population.^20^ In the US in 2021, 76% of women over age 50 report being screened within the past 2 years.^21^ Mammograms offer a unique opportunity to advance clinical prevention in parallel with the screening benefit of early diagnosis and reduced breast cancer mortality.^22^ With over 80% of women screened with DBT^10^ it is imperative that risk models now incorporate this imaging modality. Furthermore, many clinical services have stopped FFDM screening to reduce radiation exposure for women, increasing the imperative for prediction based on DBT alone.

Research to improve breast cancer risk prediction beyond the Gail model^23^ has included established risk factors, hormones, mammographic density, and polygenic risk scores as summarized a comprehensive reviews.^1^ These models based on varying combinations of demographic risk factors with addition of polygenic risk scores and mammographic breast density perform modestly at best with AUCs of 0.54 to 0.68 for prediction of 5 year risk.^24^ Advances in AI methods have also been applied to FFDM with comparable performance when using a baseline mammogram,^25,26^ and relying less on clinical factors. With broad population access to and adherence with screening recommendations and the uptake of DBT to over 80% of breast screening.^10^ Furthermore, participation in breast screening is a goal for race-ethnic groups so models must perform across the diverse US population. The need for the models to be evaluated in diverse populations and now include DBT is pressing.^6^ To date, only one evaluation of DBT for risk prediction has been identified.^2^ That study limits prediction to a 2-year time horizon with only internal validation, which is shorter than the 5-year horizon and lacks the external validation used to guide clinical practice. We overcome this methodologic gap and use a diverse external validation data source from an urban Atlanta clinical service where 46% of women are Non-Hispanic Black.^13^

Importantly, risk management guidelines move beyond the USPSTF recommendation for routine screening for women 40 to 74, to identifying women at elevated risk who may be offered genetic risk testing, risk reduction approaches (NCCN and ASCO)^3,4^ or modified screening strategies as defined by the American College of Radiology^5^. While the US uses a 5-year time horizon, in the UK a 10-year horizon is used.^27^ We note that applying the TC model in our population had relatively modest performance separating high vs low risk as seen in other settings. In other studies, it has identified a high risk population that gives rise to some 39% of cancers.^28^ Many of the demographic models use risk factors that have been shown to modify breast density,^29^ which itself is summarize or contained in the whole breast image. Thus, these crude ways to estimate the image and breast cancer risk are not contributing to estimation once the whole image is incorporated into the prediction model. For clinical translation, our calibration shows good agreement between predicted and observed and identifies 15% of the population at high risk over the next 5 years from whom 49% of total cases are diagnosed.

While the sample size for our study is somewhat limited by the relatively recent widespread use of DBT for breast cancer screening,^10^ the performance and calibration offer opportunities to apply this approach to risk prediction in routine clinical settings, consistent with current US risk management guidelines. Further evaluation of performance across additional race ethnicity groups will strengthen the applicability of this approach.

This study has important strengths. This study uses a diverse external validation population that is drawn from two community hospitals, a large inner-city hospital and a private academic hospital. Our 5-years risk model is calibrated to SEER which accommodates the use of accepted risk management cut points and allows direct application for routine clinical risks management across clinical settings. We limit the analysis to only use of synthetic DBT and begin follow-up of women free from breast cancer from that first DBT exam.

Our results should be considered in the context of limitations. Additional race-ethnic diversity of the screened populations will strengthen applicability and address gaps in evidence.^6^ Reflecting the longer use of DBT in routine screening, additional long term follow-up will become available and should provide further evidence on utility in routine care.

## Conclusions

Our 5-year risk model using synthetic DBTs accurately classifies women according to absolute risk of breast cancer and can facilitate guideline driven risk management and support more equitable screening programs.

## Data Availability

Development data mammogram images and covariates at WashU - available with data use agreement
External validation data from Emory publicly available

https://github.com/Emory-HITI/EMBED_Open_Data

